# Peripheral optical coherence tomography assisted by scleral depression in retinopathy of prematurity

**DOI:** 10.1101/2021.10.14.21265026

**Authors:** Brittni A. Scruggs, Shuibin Ni, Thanh-Tin P. Nguyen, Susan Ostmo, Michael F. Chiang, Yali Jia, David Huang, Yifan Jian, J. Peter Campbell

## Abstract

**Objective:** To determine whether handheld widefield optical coherence tomography (OCT) can be used to document retinopathy of prematurity (ROP) stage while using scleral depression to improve peripheral views.

**Design:** Prospective observational study

**Participants:** Consecutive neonates admitted to the neonatal intensive care unit (NICU) in a single academic medical center who also met criteria for ROP screening and consented for research imaging.

**Methods:** Scleral depression was combined with widefield OCT using an investigational 400-kHz, 55-degree field of view handheld OCT during ROP screening from October 28, 2020 to March 03, 2021.

**Main Outcome Measures:** Acquisition of *en face* and B-scan imaging of the peripheral retina to objectively assess early vitreoretinal pathology, including the demarcation between vascularized and anterior avascular retina, the presence of early ridge formation, and small neovascular tufts.

**Results:** Various stages of ROP were detected using a rapid acquisition OCT system. In one neonate, serial OCT imaging over a five-week period demonstrated accumulation of neovascular tufts with progression to stage 3 ROP with extraretinal fibrovascular proliferation along the ridge. Videography of this technique is included in this report for instructional purposes.

**Conclusions:** Serial examinations using widefield OCT and scleral depression may improve detection and documentation of ROP disease progression. Earlier detection of ROP-related proliferation may prevent vitreoretinal traction, retinal detachment, and blindness.

## INTRODUCTION

The routine use of optical coherence tomography (OCT) in retinopathy of prematurity (ROP) has been limited, despite the advantages OCT provides in the care of patients with retinal diseases. In adults, the leading causes of blindness include age-related macular degeneration and diabetic retinopathy; in both, OCT is more sensitive than the clinical exam for detection of severe disease, and it has become standard of care and facilitated the transition towards quantitative and objective disease screening, diagnosis, and monitoring. This has resulted in opportunities for earlier treatment and improved visual outcomes.

In ROP, the standard of care remains the ophthalmoscopic exam, with subjective assessment of zone, stage, and plus disease. Prior applications of OCT in ROP have been limited by the field of view (FOV) of the commercially available and investigational OCT systems. Thus, most early applications of OCT have focused on posterior manifestations of ROP, rather than assessment of peripheral ROP stage.^1-7^ Previous work has demonstrated that OCT may provide added information about foveal architecture,^2^ vitreous organization,^6^ three dimensional manifestations of plus disease,^7^ and other clinical signs that we cannot appreciate using the clinical exam. In addition, Mangalesh, *et al*. recently demonstrated the potential for reduced neonatal stress using OCT rather than the clinical exam, which is a key potential benefit of screening with OCT.^1^

Unlike in most applications of OCT, the primary pathology in ROP is peripheral; therefore, the most compelling case would be made if OCT could reliably diagnose the stage of disease, potentially replacing the ophthalmoscopic examination. Several groups have demonstrated investigational OCT devices can achieve wider fields of view, roughly equivalent to the visualization provided by indirect ophthalmoscopy; however, these still cannot routinely visualize the retinal periphery.^3,8^ Given that clinical assessment of peripheral ROP with ophthalmoscopy often requires scleral depression, we tested in this study the hypothesis that scleral depression, combined with the use of a widefield OCT system, could provide accurate, objective, and rapid assessment of ROP stage.

## METHODS

Handheld OCT retinal imaging was performed on fully awake neonates in the neonatal intensive care unit (NICU) at Oregon Health & Science University (OHSU) during the period of October 28, 2020 to March 03, 2021. Infants were included in the study if they met eligibility criteria for ROP screening (birth weight ≤1500g and/or a gestational age ≤30 weeks), and if parent(s) consented for research imaging. Any infant whose parent(s) declined participation in the study were excluded and did not undergo research imaging; however, those infants still underwent ROP screening using traditional methods, including indirect ophthalmoscopy. Thirteen infants were included in this study, and the research OCT images were obtained at each visit during usual ROP screening. Birthweight, gestational age, gender, race/ethnicity, ROP zone by clinical assessment, and ROP stage by clinical assessment were recorded for all infants (Table 1). The imaging for the non-sedated infants was conducted with pharmacological dilation with cyclopentolate 0.2% and phenylephrine 1%. After installation of anesthetic eye drops, an infant lid speculum was utilized, scleral depression was performed in a manner similar to the clinical ROP examination. This study was approved by the Institutional Review Board/Ethics Committee of OHSU in accordance with the Declaration of Helsinki.

An investigational 400-kHz, 55-degree FOV portable handheld OCT system was used in this study.^9^ This portable handheld OCT extends views to the peripheral retina of pediatric patients and displays cross-sectional (*i*.*e*., B-scan) and *en face* views in real time.^10^ With a single hand, the operator could hold the lens tube, allowing the operator to rest parts of their palm gently on the infant’s forehead and to manipulate the eye position using a depressor with their other hand (Video 1). The acquisition time of each volume was 120 ms with 400 A-scan per B-scan and 120 B-scans per volume, which provided real-time feedback in optimizing images. Once the target area on the ocular fundus was located, autofocusing with an electronically focus tunable lens could be performed in 1 second based on the brightness of the *en face* view.

The OCT images were reviewed in real-time by the examiners (S.O., Y.J., and J.P.C.), and additional images were taken, when needed, to obtain good quality images of the retinal periphery for each infant, except when clinical needs of the baby superseded the value of obtaining research imaging. OCT images were obtained of the temporal, posterior, and nasal retina at every session, as well as the superior and inferior when relevant pathology was suspected or noted. The ROP zone and stage associated with any OCT image were determined by the examiner using traditional examination with indirect ophthalmoscopy. RetCam imaging was performed in select cases to document clinical findings.

## RESULTS

In this study, 13 awake neonates in the NICU were imaged during routine ROP screening with serial imaging during the study period when possible. Imaging was obtained in nearly every eligible patient, excepting those whose clinical instability prevented the acquisition of research images (fewer than 3 babies during study period). In total, forty clinical visits were completed, and OCT images were obtained at each session to view the retinal periphery of each infant. Using the *en face* OCT view of the periphery with scleral depression, the extent of vascularization (zone) and any vascular abnormalities (stage) could be determined, as shown in Figure 1 and Video 2. In eyes that were fully vascularized, the scleral depressor could be visualized with retinal vessels extending within one disc area of the ora serrata (Video 1). In contrast, in eyes that demonstrated persistent avascular retina, or any degree of stage, the edge of the vascularized retina could be viewed *en face* (Figure 1 & Video 2).

**Figure 1.**
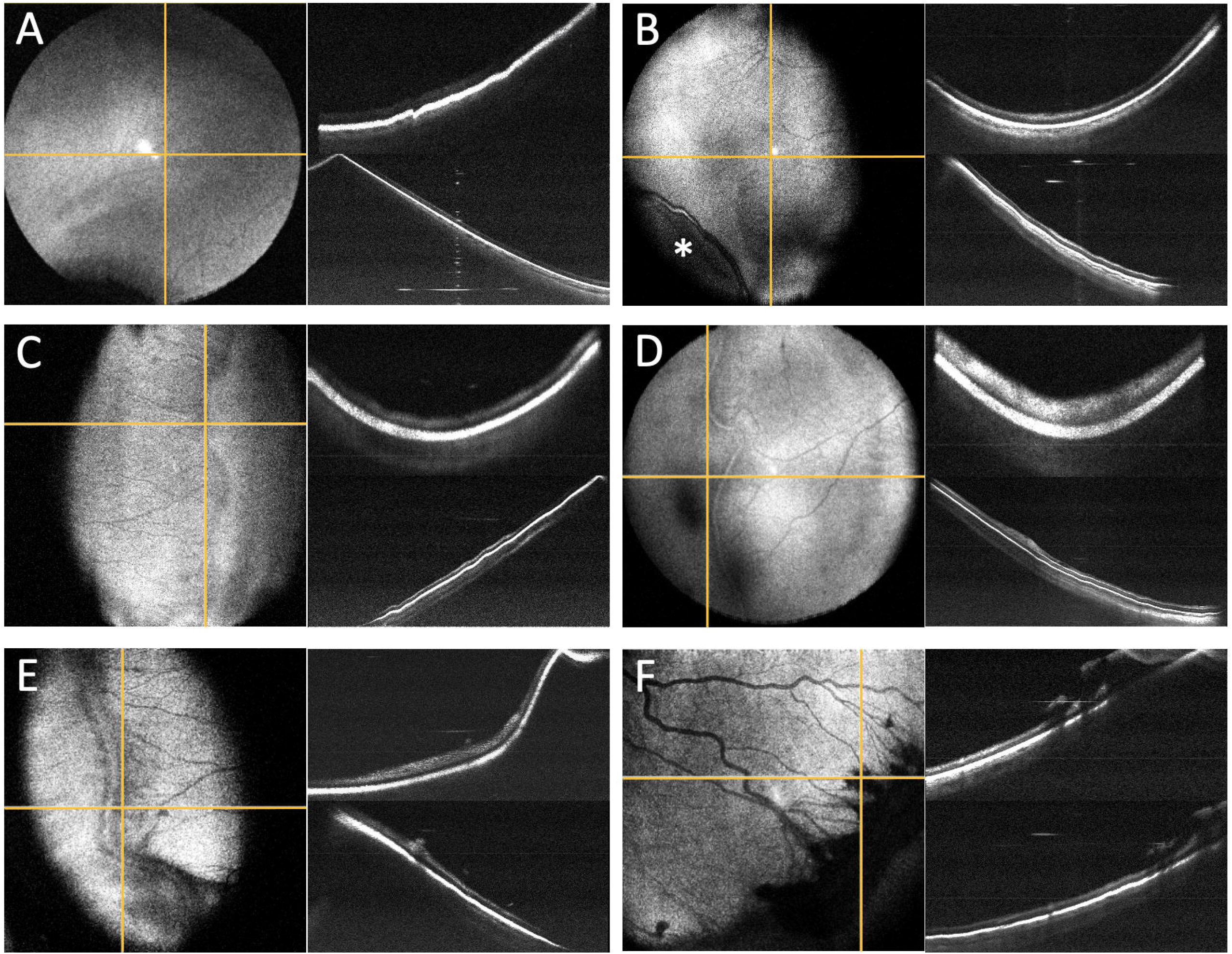
ROP peripheral pathology as seen with peripheral OCT *en face* and cross-sectional imaging using scleral depression. A, Avascular retina without clear vascular-avascular border (sometimes referred to as stage 0 ROP). B-C, Two examples of stage 1 ROP visible with faint (B) and more pronounced (C) demarcation lines without ridge formation. The asterisk indicates scleral depressor location. D-E, Two examples of stage 2 ROP with early (D) and later (E) ridge formation without neovascularization. F, Stage 3 ROP with significant extraretinal fibrovascular proliferation along the ridge. The yellow vertical lines correlate to the upper cross-sectional images; yellow horizontal lines correlate to lower cross-sectional images.

We observed a spectrum of vascular abnormalities ranging from avascular retina without a clear demarcation line (sometimes referred to as stage 0; Fig 1A), to extraretinal neovascularization (stage 3; Fig 1F). Even when comparing two neonates with clinical stage 1 ROP (Fig 1 B-C), there were meaningful differences in OCT appearance, including B-scan images. Fig 1B shows a faint demarcation line separating the vascularized and anterior avascular retina without dimension, whereas Fig. 1C shows a more distinct demarcation line. Similarly, the images in Fig 1 D-E show early stage 2 ROP (Fig 1D) compared to the prominent, more vascularized later stage 2 ROP (Fig 1E).

This technique can also be used to monitor the progression of ROP over time. Figure 2 and Video 3 show OCT *en face* images of the posterior pole and peripheral retina obtained with B-scan images of the peripheral pathology in one neonate over a five-week period. This one example demonstrates two important concepts. First, while the anterior ridge slowly progressed into a typical stage 3 lesion by week 5, the serial OCT imaging demonstrated accumulation of “popcorn” neovascular tufts (typically associated with stage 2 ROP) with clear and continuous progression of extraretinal fibrovascular proliferation along the ridge in the weeks prior. Second, the *en face* images centered on the posterior pole showed increased vascular dilation and tortuosity over time, correlating with the increased severity of peripheral disease.

**Figure 2.**
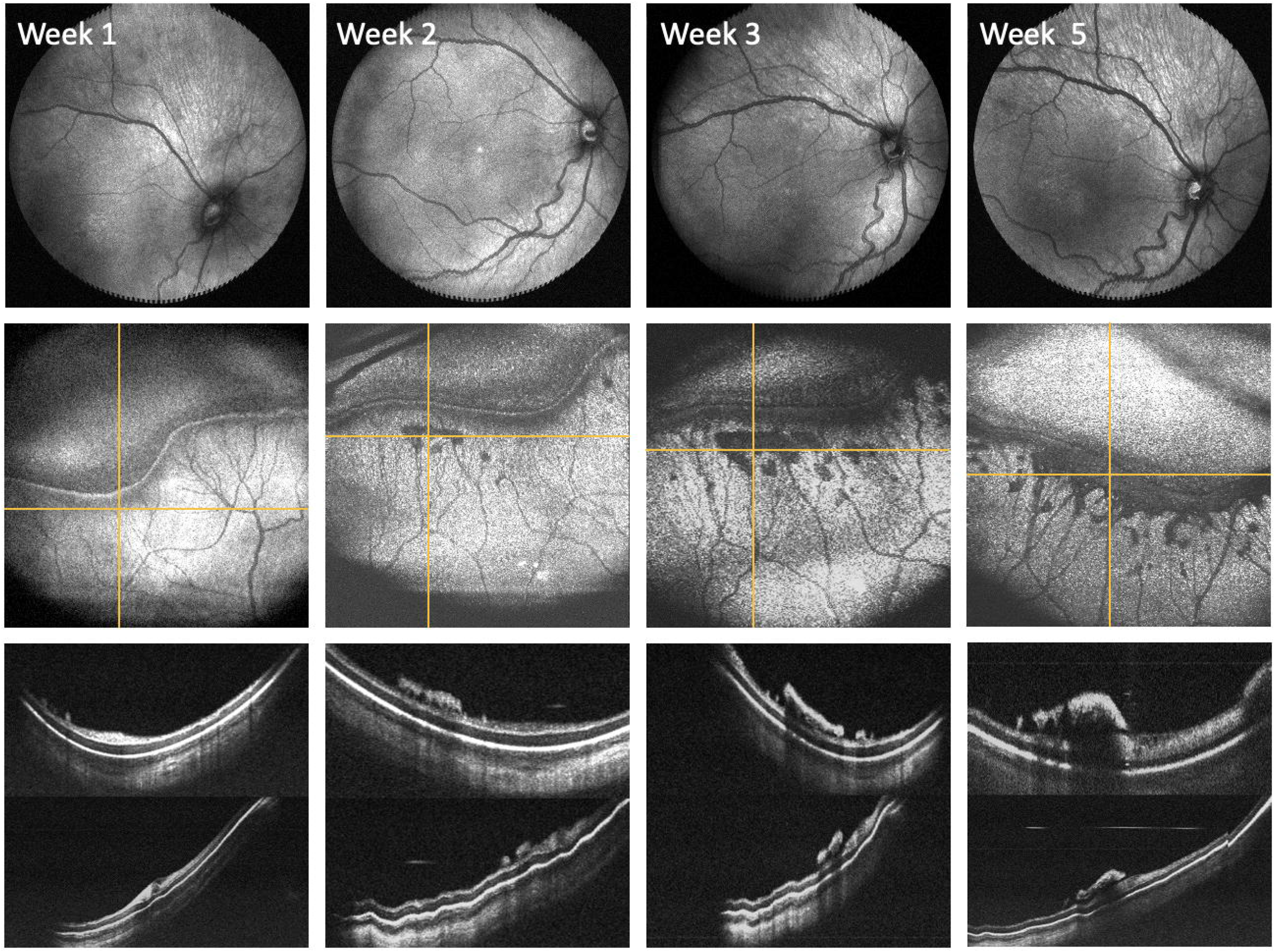
Monitoring ROP disease progression in a preterm infant using OCT *en face* and cross-sectional views. **Top row:** Posterior *en face* views over a five-week period demonstrate increased vascular tortuosity. **Second row:** Serial peripheral *en face* views centered on the patient’s peripheral ridge show increased severity of ROP disease over time. The yellow horizontal lines correlate to the respective cross-sectional images obtained posterior to the ridge (**Third row, top image**). The yellow vertical lines correlate to the respective cross-sectional images obtained across the ridge (**Third row, bottom image**).

## DISCUSSION

In this report, we demonstrate that the peripheral ROP stage can be documented with OCT using scleral depression and that stage appears to represent a spectrum from avascular retina to extraretinal neovascularization. Zone III is clearly visualized using technique, and these far peripheral views provide objective documentation of the extent of vascularization and/or avascular retina for preterm infants. This study also suggests that changes in the posterior pole in the spectrum of pre-plus and plus disease correspond to the degree and progression of stage, as has been seen with artificial intelligence-based assessment of vascular severity, but further work is needed to establish the relationship via OCT. ^11^

Videography of this entire process has been provided to demonstrate the utility of this imaging modality at bedside with minimal disruption to the neonate or ROP screening process. We have found that incorporating OCT in our practice, especially when performed with scleral depression, improves our ability to visualize peripheral ROP, to teach trainees, to communicate findings with NICU staff and parents, and to visualize early vitreoretinal traction not visible with ophthalmoscopy.

Using the *en face* real-time viewing, OCT can be used as an ophthalmoscope. In addition to visualizing the vasculature, OCT can objectively assess the degree of vascular abnormalities. Chen, *et al* previously demonstrated OCT examples at the vascular-avascular junction with comparison to prior histology.^3^ In this report, we demonstrate that with scleral depression it may be feasible to utilize OCT in ROP practice, overcoming one of the limitations to its use. Although in this series we did not observe any retinal detachments, an added advantage of using OCT to detect extraretinal fibrovascular tissue is that it may be more sensitive for detection of “flat” extraretinal neovascularization associated with aggressive ROP (AROP) and earlier signs of vitreoretinal traction, which can be very difficult to appreciate clinically.

## CONCLUSION

In this study, the combination of widefield OCT and scleral depression allow detection and documentation of ROP stage and progression of peripheral pathology. As the cost of this technology comes down, we hope that reports like this demonstrating added clinical value may lead to routine use of OCT as part of ROP screening, earlier detection of progressive stage and extent of disease, fewer retinal detachments, and improved visual outcomes in ROP.

## Data Availability

All data produced in the present study are available upon reasonable request to the authors.

## Video Legends

**Video 1**. Visualization of the peripheral retina during ROP screening using scleral depression and OCT. E*n face* views and cross-sectional scans are shown simultaneously in real time. The scleral depressor, seen prior to image acquisition, provides indentation throughout this process and facilitates view of peripheral vasculature. This video demonstrates the rapid acquisition time of the investigational portable handheld OCT system.

**Video 2**. Documentation of various ROP stages detected by OCT using scleral depression. This video shows real time acquisition of OCT e*n face* and cross-sectional scans of five preterm infants. The images are shown in increasing severity of ROP peripheral pathology. The videography clips of Patients 1-5 correspond with the images in Fig 1 B-F, respectively. Patients 1 and 2 demonstrate examples of stage 1 ROP, whereas patients 3 and 4 had clinical evidence of stage 2 ROP. Patient 5 had extraretinal fibrovascular proliferation, consistent with stage 3 ROP.

**Video 3**. OCT detection of peripheral ROP disease progression. OCT *en face* views centered on a patient’s peripheral ridge demonstrate increased severity of ROP disease over a five-week period. The adjacent cross-sectional images demonstrate an increase in “popcorn” neovascularization tufts and extraretinal fibrovascular tissue over this period. By week 5, the patient had evidence of stage 3 ROP and underwent treatment.

